# The effectiveness of the three-tier system of local restrictions for control of COVID-19

**DOI:** 10.1101/2020.11.22.20236422

**Authors:** Paul R Hunter, Julii Brainard, Alastair Grant

## Abstract

Despite it being over 10 months since COVID-19 was first reported to the world and it having caused over 1.3 million deaths it is still uncertain how the virus can be controlled whilst minimising the negative impacts on society and the economy. On the 14^th^ October, England introduced a three-tier system of regional restrictions in an attempt to control the epidemic. This lasted until the 5^th^ November when a new national lockdown was imposed. Tier 1 was the least and Tier 3 the most restrictive tiers. We used publicly available data of daily cases by local authority (local government areas) and estimated the reproductive rate (R value) of the epidemic over the previous 14 days at various time points after the imposition of the tier system or where local authorities were moved into higher tiers at time points after reallocation. At day 0 there vas very little difference in the R value between authorities in the different groups but by day 14 the R value in Tier 3 authorities had fallen to about 0.9, in Tier 2 to about 1.0 and in Tier 1 the R value was about 1.5. The restrictions in Tier 1 had little impact on transmission and allowed exponential growth in the large majority of authorities. By contrast the epidemic was declining in most Tier 3 authorities. In Tier 2, exponential growth was being seen in about half of authorities but declining in half. We concluded that the existing three tier system would have been sufficient to control the epidemic if all authorities had been moved out of Tier 1 into tier 2 and there had been more rapid identification and transfer of those authorities where the epidemic was increasing out of Tier 2 into Tier 3. A more restrictive tier than Tier 3 may be needed but only by a small number of authorities.

## Introduction

Since the end of 2019 the SARS-CoV-2 virus has spread globally and caused over 55 million cases and more than one million deaths from COVID-19. Several publications in recent months attempted to demonstrate what nonpharmaceutical interventions may or may not be successful in controlling the pandemic (Brauner *et al*. 2020, Hunter *et al*. 2020). There has also been debate as to the value of regional versus national control strategies. In October 2020 England introduced a three tier system for controlling COVID-19 (Prime Minister’s Office 2020), with social mixing restrictions ranging from limits on maximum group sizes in Tier 1 through to a ban on household mixing indoors or in private gardens in Tier 3. The system was meant to be flexible with all placements subject to ongoing review. Then on the 5th of November the English government imposed a national “lockdown” in which people were instructed to stay at home except for specific purposes; non-essential retail was closed, but schools and universities remained open (Department of Health and Social Care 2020). That a lockdown was about to be imposed was widely rumoured in news media prompting what appeared to be an unplanned announcement about the national lockdown about five days before it came into force (Blackall 2020). At the time of writing, the national lockdown in England is due to finish in on 2 December. It has been suggested that England will revert to a multi-tier control strategy similar to what was implemented in October. It would therefore be very valuable to understand whether or not the English three tier system was managing to control the epidemic prior to the imposition of the “lockdown”. This paper presents a review of daily reported statistics of new infections by local authority in England and seeks to distinguish the transmissibility of COVID-19 depending on which tier local authorities had been placed.

## Methods

All data on daily numbers of new cases of COVID-19 were downloaded from the English Department of Health and social care daily COVID dashboard. Daily data by local authority is maintained at https://coronavirus.data.gov.uk/details/about-data.

For each local authority, an estimate of the effective reproductive number (R) for preceding days was obtained by summing all reports of new COVID infections for each day and the previous six days. This was then compared with the sum of new cases over the previous seven-day period. R was estimated using the equation given by Grant (2020) as

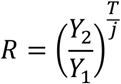

Where Y_1_ and Y_2_ are the numbers of cases in two consecutive weeks, T is the generation time for the infection (here taken as 5 days) and j is the interval between the midpoints of the periods over which cases are counted (here 7 days). This estimator performs particularly well when R is close to 1 (Grant 2020).

In order to demonstrate the impact of the different tiers we estimated R values for all local authorities for days 0, 7, 14, 21 and 28 where day zero was the day that the three tier system was implemented (starting 14^th^ October). For those authorities that remained in Tier 1 throughout the four weeks the dates were the same. For local authorities that were in Tiers 2 or 3 on the 4^th^ November day 0 was not the 14^th^ October but rather the day that they were placed into their final tier and all subsequent dates taken from then. Local authorities that moved up a tier were not included in any analyses of their initial tier group. Instead any analyses from local authorities that started in one tier but moved into a higher tier were included for the lower tier up to 7 days after the reallocation of tier. Analyses in the newer higher tier were included as above.

The mean and standard deviations of the R values for each local authority were calculated in Microsoft Excel. The distribution of R values for local authorities in their final allocation is presented in box and whisker plots. Data from dates up to the 12^th^ of November were analysed, seven days after implementation of the national lockdown. COVID-19 has a median incubation period of about 5 days (Lauer *et al*. 2020), and test results tend to take 24-48 hours to return (Department of Health and Social Care 2020). Therefore, it was felt that data after 12 November, the restrictions associated with national lockdown rather than tiers would mostly influenced case counts. Comparison of the counts in different tiers was done using one way analysis of variance.

## Results

Data were obtained on 315 local authorities. One local authority, High Peak (in East Midlands), was excluded from the analyses for Tier 2 because at the start only parts of High Peak were placed in Tier 2 to be followed days later by the remainder of the authority area.

Figure 1 shows the daily numbers of new cases reported in England based on the swab sample collection date. It can be seen that case numbers increased rapidly from early September through to 5 October, after which there was a less rapid increase till 16 October and then much slower increase from about the 23^rd^ to 30^th^ October, after which case numbers accelerated once more.

**Figure 1.**
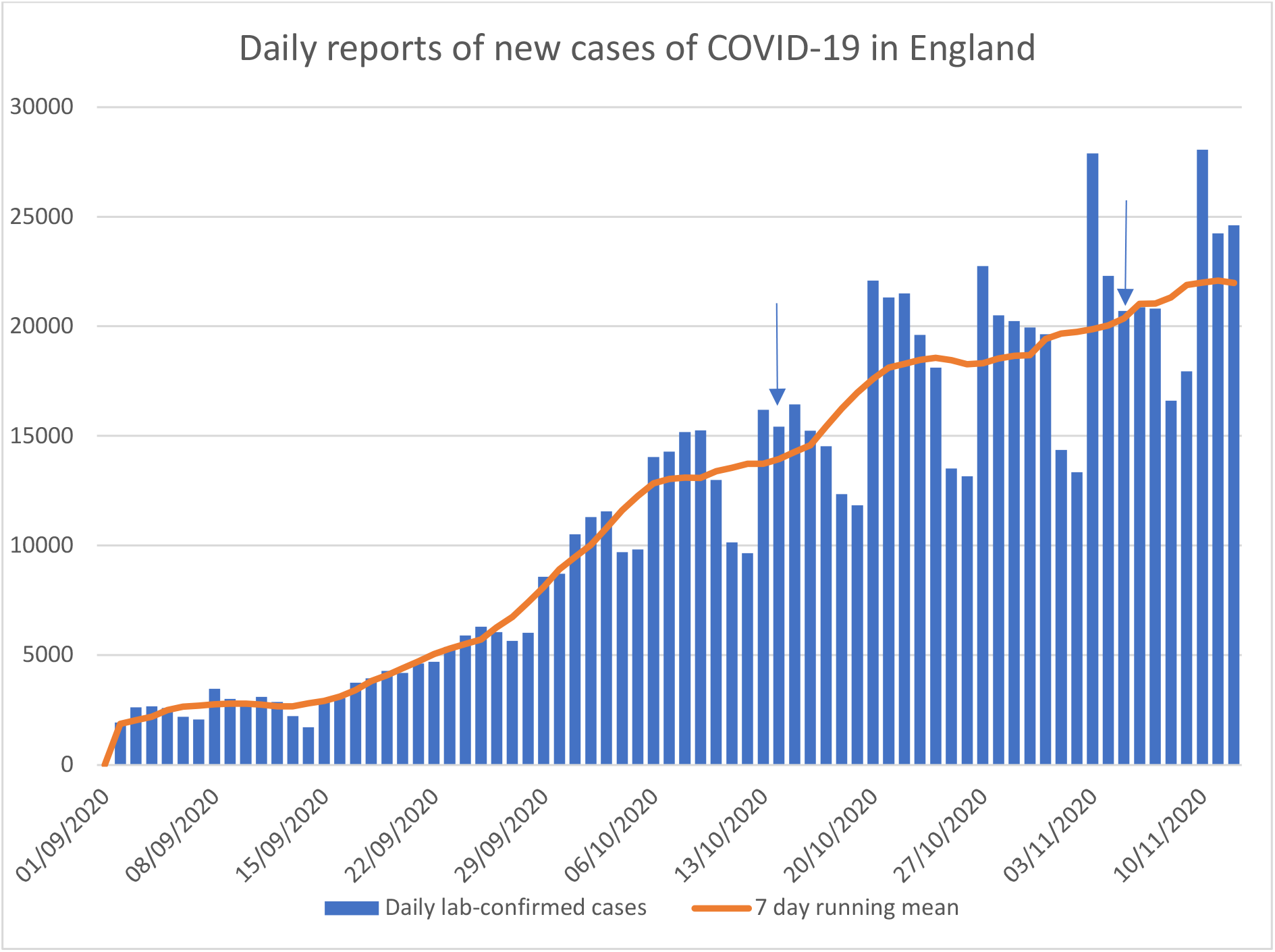
Daily reports of new cases of COVID-19 in England based on sample collection date.^a^. ^a^ Arrows indicate start of tier system on 14^th^ October and start of national lockdown on 5^th^ November

Table 1 shows the mean and standard deviation of the estimated R for the authorities combined and disaggregated by allocated tier. It can be seen that for all included authorities, R prior to the start of implementation was above 1.1, indicating exponential growth. As discussed above, the R value was calculated giving an estimate over the previous 14 days. During the following four weeks the mean R value remained over 1.1. In Tier 1 the R values also remained fairly constant throughout the four weeks, indicating (at least in Tier 1) continued exponential growth. For Tier 2, on the date of implementation R was similar to that in Tier 1. However, by day 14 the R value had fallen to about 1.0, though on day 28 there was evidence that infection transmission had started increasing over the previous week. For Tier 3 authorities on day zero the R value was similar to the other tiers but by day 14, R had fallen to about 0.9 where it remained. It can also be seen that as the R value in those initial

**Table 1.** Summary statistics for the R values from each of the Local Authorities by tier

|  |  | Days after implementation |  |  |  |  |
| --- | --- | --- | --- | --- | --- | --- |
|  |  | Day 0 | Day 7 | Day 14 | Day 21 | Day 28 |
| All local authorities | Mean | 1.15 | 1.22 | 1.11 | 1.10 | 1.15 |
|  | Std Dev | 0.20 | 0.20 | 0.19 | 0.18 | 0.20 |
|  | N | 315 | 315 | 315 | 315 | 315 |
| Authorities remaining in Tier 1 throughout | Mean | 1.15 | 1.22 | 1.16 | 1.14 | 1.20 |
|  | Std Dev | 0.23 | 0.22 | 0.20 | 0.19 | 0.21 |
|  | N | 169 | 169 | 169 | 169 | 169 |
| Authorities in Tier 1 prior to moving into Tier 2 | Mean | 1.18 | 1.36 | 1.21 | 1.18 |  |
|  | Std Dev | 0.16 | 0.18 | 0.16 | 0.15 |  |
|  | N | 76 | 26 | 26 | 22 | 0 |
| Authorities in Tier 2 on 4th November | Mean | 1.13 | 1.19 | 0.98 | 1.06 | 1.17 |
|  | Std Dev | 0.13 | 0.16 | 0.13 | 0.13 | 0.07 |
|  | N | 97 | 97 | 74 | 71 | 21 |
| Authorities in Tier 2 prior to moving into Tier 3 | Mean | 1.14 | 1.20 | 1.05 | 1.04 |  |
|  | Std Dev | 0.18 | 0.17 | 0.09 | 0.10 |  |
|  | N | 44 | 28 | 28 | 13 | 0 |
| Authorities in Tier 3 on 4th November | Mean | 1.13 | 1.04 | 0.89 | 0.92 | 0.94 |
|  | Std Dev | 0.15 | 0.12 | 0.07 | 0.09 | 0.09 |
|  | N | 48 | 47 | 34 | 19 | 6 |
| One way ANOVA comparing R values between Tiers 1, 2 and 3 | F (variance ratio) | 0.49 | 16.63 | 50.80 | 17.22 | 5.36 |
|  | P | 0.613 | <0.00001 | <0.00001 | <0.00001 | 0.005 |

\*Excluding High Peak from Tier 2 analyses as discussed in the text.

Tier 1 authorities that were moved up to Tier 2 during the time period had higher R values than those Tier 1 authorities but that there was little difference between those Tier 2 authorities that remained in Tier 2 and those that were eventually moved up to Tier 3 prior to their move.

A one way analysis of variance comparing the mean R values in the Local Authorities in the three tiers showed no difference on day 0 but a very highly significant difference on days 7, 14 and 21, especially days 14 and 21. On day 28 the difference was still significant but much less so, but this primarily reflects the relatively small number of authorities that could be included in day 28.

Figure 2 shows the box and whisker plots for all authorities and for each of their final tiers. The large majority of local authorities in Tier 1 had R values >1 and remained in exponential growth throughout the 28 days. By contrast, the majority of Tier 3 authorities had R < 1 at least by Day 14 and so in these authorities the epidemic was in decline. Tier 2 shows an intermediate pattern, with about half of authorities continuing to see exponential growth on Day 14 while the remainder saw some decline, but the median value of R in Tier 2 authorities then increased on Days 21 and 28

**Figure 2.**
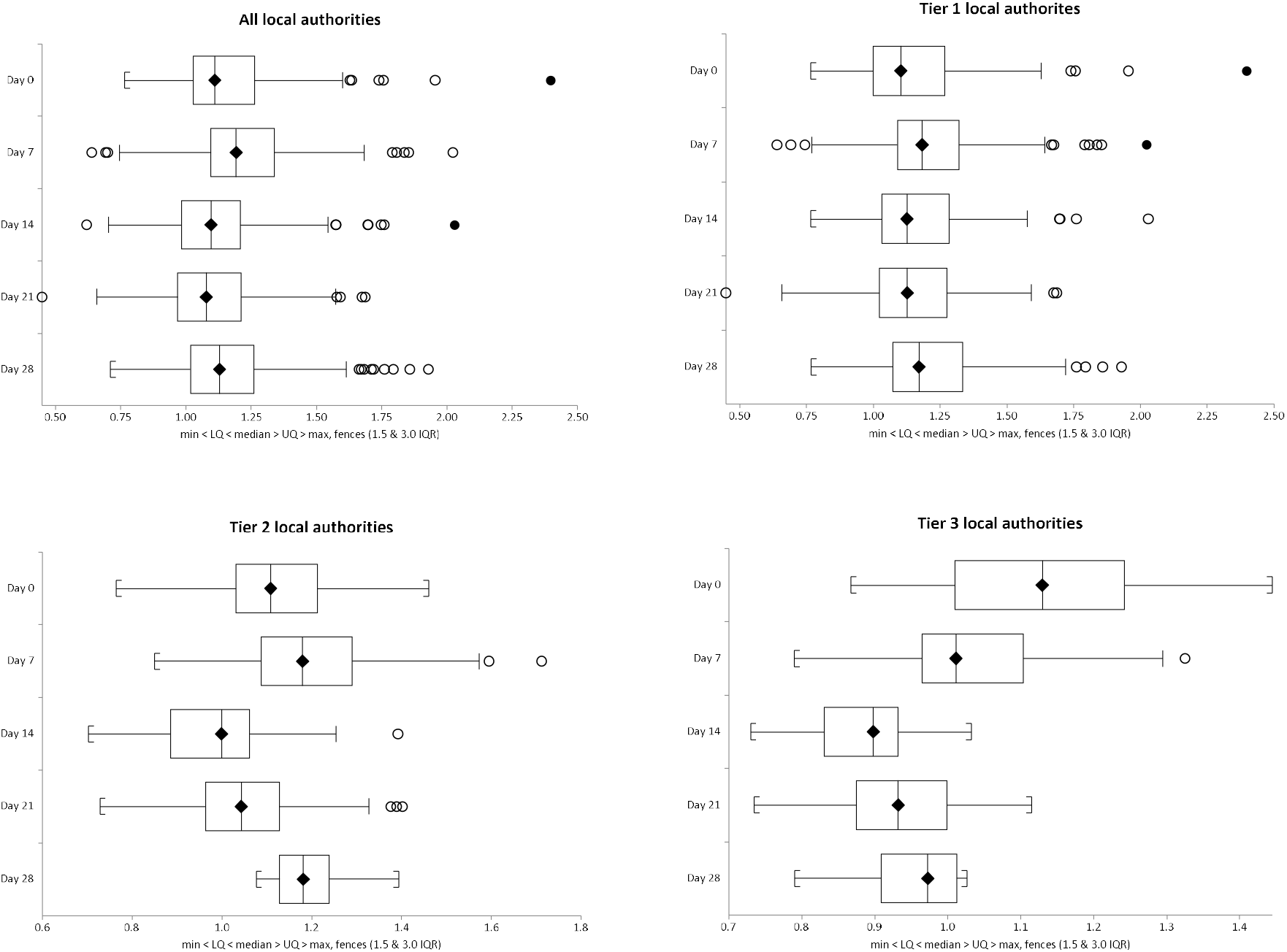
Box and Whisker plots of distribution of estimated of R values by days after implementation of the tier system or for Tiers 2 and 3 by days after movement into these final tiers by local authority combined and in different tiers based on final tier allocation on 4^th^ November.

## Discussion

We undertook an analysis of new cases of COVID-19 by local authority in England from the 14th of October to the 12th of November. We demonstrated that the epidemic continued to grow in most areas during this time. We further showed that the rate of growth of the epidemic could largely be predicted by which tier a local authority was placed in. In Tier 1, the least restricted tier, the large majority of authorities continued to see exponential growth. This is in contrast to Tier 3, where most authorities saw a decline in cases during the same time and a there was a reduction in the R values to less than 1. Analysis of Tier 2 authorities found that that overall case numbers remained static after the first week or so. Roughly half of the authorities in Tier 2 continued to see some growth and the other half had decline. Of note is that for Tier 2 there was little difference in the R value between authorities that were moved from Tier 2 into 3 and those that remained in Tier 2. This may suggest that the how rapidly a local epidemic was growing had little effect on the decision about whether authorities were placed in Tiers 2 or 3.

This is a retrospective observational study and, as such is subject to potential confounding. However, we are not aware of events that could have affect the epidemic other than which tier the local authority had been placed in. The data from day 28 slightly overlap with the national lockdown period which started 5th of November. However, it is unlikely that in the first 7 days of the national lockdown there would have been much impact on case numbers given the time to incubate the disease and then obtain a test result. On the other hand, case numbers were noted to increase more rapidly in early November than they did in the last week of October. It has been suggested that this increase in cases in the early November may have been due to increased socialisation after the national lockdown was announced and before it was implemented. At present, this has yet to be proven. We did note some increases in R values at day 28 in all tiers so need to be cautious about interpreting the impact of the tier system at that time point.

Whilst our analysis shows a clear difference in R values between local authorities depending on which tier they are in, it cannot be determined from our analysis what aspects of the restrictions in Tiers 1-3 were responsible for case count outcomes. In the UK, Bernal *et al*. (2020) reported that secondary transmission within households is one of the most important risk factors for acquiring the infection. Nevertheless, banning gatherings of groups of people have been shown to be associated with reduced transmission even when restricting gatherings to less than 10 (Brauner *et al*. 2020, Hunter *et al*. 2020; Li *et al*, 2020; Cevik et al. 2020). One of the main characteristics of the English Tier system was in fact to reduce the opportunities for groups of people to gather indoors in any number. It is believed that “superspreaders” who infect large numbers of other individuals, are crucial to the continued spread of the virus (Althouse *et al*. 2020) and it is likely that superspreading events would also have been less like given the reduced opportunities for gatherings with the higher English Tiers.

In general, higher tiers allowed less social interaction between people from different households, especially in enclosed environments. For instance, Tier 1 allowed for groups of maximum six persons from different households to meet indoors or outdoors. Tier 2 permitted such groups to meet only outdoors up to six people. In Tier 3 people were not allowed to socialise with anyone they did not live with indoors or outdoors in private gardens or most outdoor hospitality venues unless they were part of mutual support ‘bubbles’. Groups of up to six were allowed to meet in outdoor public spaces such as parks. There were corresponding increased social mixing restrictions described by the Tier system with respect to many other activities, such as sport, rules while visiting restaurants, travel outside home area, etc. Consequently our findings are consistent with what would be expected from previous studies (Brauner *et al*. 2020, Hunter *et al*. 2020; Li *et al*, 2020; Cevik et al. 2020).

In conclusion, we have shown that the tier system probably was helping to reduce transmission of the epidemic in England. However, case numbers were continuing to increase albeit more slowly than previously. Placement in Tier 1 was clearly inadequate to control spread for most authorities, while Tier 3 was highly effective in the great majority of local authorities. In around half of the local authorities in Tier 2 were still seeing exponential growth.

It seems likely that the tier system will better prevent spread of SARS-CoV-2 with faster identification of the appropriate placement of each local authority and swifter reallocation when needed. Our findings would suggest that about half of authorities originally placed in Tier 2 would more appropriately have been placed in tier 3 and almost all of the authorities originally placed in Tier 1 should have been placed in Tier 2. It has yet to be decided how England will move forward when the current national lockdown comes to an end. Whilst an additional even more restrictive tier may be needed, the evidence from Tier 3 areas so far suggests that Tier 3 restrictions are likely to adequate for most but not necessarily all local authority areas. What is needed as we move out of the national lockdown is a faster mechanism to identify those areas where the epidemic is not being controlled that can enable more-timely placement into a more appropriate tier. However, the extent to which there is social acceptance of, and compliance with, restrictions is at least as important as the level of restrictions that are nominally in force (Bellato 2020). Self-reported compliance with many of the social contact rules implemented to prevent virus spread most of the time has been fairly good, but not perfect in the UK (Bellato, 2020; Office for National Statistics 2020). However, there is evidence of widespread confusion about social distancing regulations and when a SARS-CoV-2 test is appropriate, impacting on people’s ability to comply with them (Office for National Statistics 2020; Williams et al 2020; Smith et al 2020). No formal set of guidelines and restrictions will be effective unless the general public understand and support the restrictions and control strategy.

## Supporting information

STROBE checlist

## Data Availability

All data is publicly available from the UK Department of Health and Social Care COVID-19 Dashboard

https://coronavirus.data.gov.uk/details/about-data

